# A single dose of SARS-CoV-2 FINLAY-FR-1A dimeric-RBD recombinant vaccine enhances neutralization response in COVID-19 convalescents, with excellent safety profile. A preliminary report of an open-label phase 1 clinical trial

**DOI:** 10.1101/2021.02.22.21252091

**Authors:** Arturo Chang-Monteagudo, Rolando Ochoa-Azze, Yanet Climent-Ruiz, Consuelo Macías-Abraham, Laura Rodríguez-Noda, Carmen Valenzuela-Silva, Belinda Sánchez-Ramírez, Rocmira Perez-Nicado, Raúl González-Mugica, Tays Hernández-García, Ivette Orosa-Vázquez, Marianniz Díaz-Hernández, María de los A. García-García, Yanet Jerez-Barceló, Yenisey Triana-Marrero, Laura Ruiz-Villegas, Luis Rodríguez-Prieto, Rinaldo Puga-Gómez, Pedro Pablo Guerra-Chaviano, Yaíma Zúñiga-Rosales, Beatriz Marcheco-Teruel, Mireida Rodríguez-Acosta, Enrique Noa-Romero, Juliet Enríquez-Puertas, Delia Porto-González, Kalet Leon-Monzon, Guan-Wu Chen, Luis Herrera Martinez, Yury Valdés-Balbín, Dagmar García-Rivera, Vicente Vérez-Bencomo, The Soberana 01B Clinical Trial team

**Author notes:** Contributed equally (Arturo Chang-Monteagudo, Rolando Ochoa-Azze, Yanet Climent-Ruiz, Laura Rodríguez-Noda).

## Abstract

We evaluated response to a single dose of the FINLAY-FR-1A recombinant dimeric-RBD base vaccine during a phase I clinical trial with 30 COVID-19 convalescents, to test its capacity for boosting natural immunity. This short report shows: a) an excellent safety profile one month after vaccination for all participants, similar to that previously found during vaccination of naïve individuals; b) a single dose of vaccine induces a >20 fold increase in antibody response one week after vaccination and remarkably 4-fold higher virus neutralization compared to the median obtained for Cuban convalescent serum panel. These preliminary results prompt initiation of a phase II trial in order to establish a general vaccination protocol for convalescents.

## Introduction

By the end of February 2021, the number of COVID-19 cases reported worldwide^1^ is reaching 111 million. COVID-19 convalescents are not included in vaccination programs and there is insufficient understanding of the efficiency and duration of protection conferred via natural immunity induced by SARS-CoV-2 infection. Is it efficient and long-lasting? Does it protect against mutant strains? What are the pros and cons of vaccinating convalescents? Do they develop adverse events not observed in the naïve population?

A vaccine based on recombinant dimeric receptor binding domain (d-RBD, 50μg) on alum (FINLAY-FR-1A) in clinical development in Cuba for protection of naïve individuals has shown an excellent safety profile.^2^ We hypothesize that a single dose of this vaccine may be an effective booster for individuals with pre-existing immunity to SARS-CoV-2. Here we describe the safety and antibody responses after application of a single dose of FINLAY-FR-1A vaccine to 30 individuals with documented pre-existing SARS-CoV-2 natural immunity. Participants were distributed in three groups: those with positive PCR test at the moment of diagnosis and cleared at least two months before the initiation of the study. Group A, who had developed mild COVID-19 disease (N=11); Group B, asymptomatic (N=10) at least two months after diagnosis; and a group C, selected from SARS-CoV-2 seropositive individuals, but who were never confirmed as PCR positive (N=9). (For a detailed description, see supplementary material).

### Vaccine associated adverse events

A concern for any vaccine, this one in convalescents of SARS-CoV-2 infection, is frequency and severity of adverse events. Individuals seropositive to SARS-CoV-2 who received one dose of an mRNA vaccine had higher frequency of adverse events compared to seronegative individuals, with at least one side effect attaining 75% of those vaccinated.^3^ With FINLAY-FR-1A, in a follow-up of adverse events over 28 days post-vaccination, only 6 individuals (20%) experienced a total of 7 vaccine associated adverse event (Figure 1), with local events predominating over systemic. This safety profile is comparable to our experience in a previous clinical trial with naïve subjects (N=20; unpublished data).^1^

**Figure 1.**
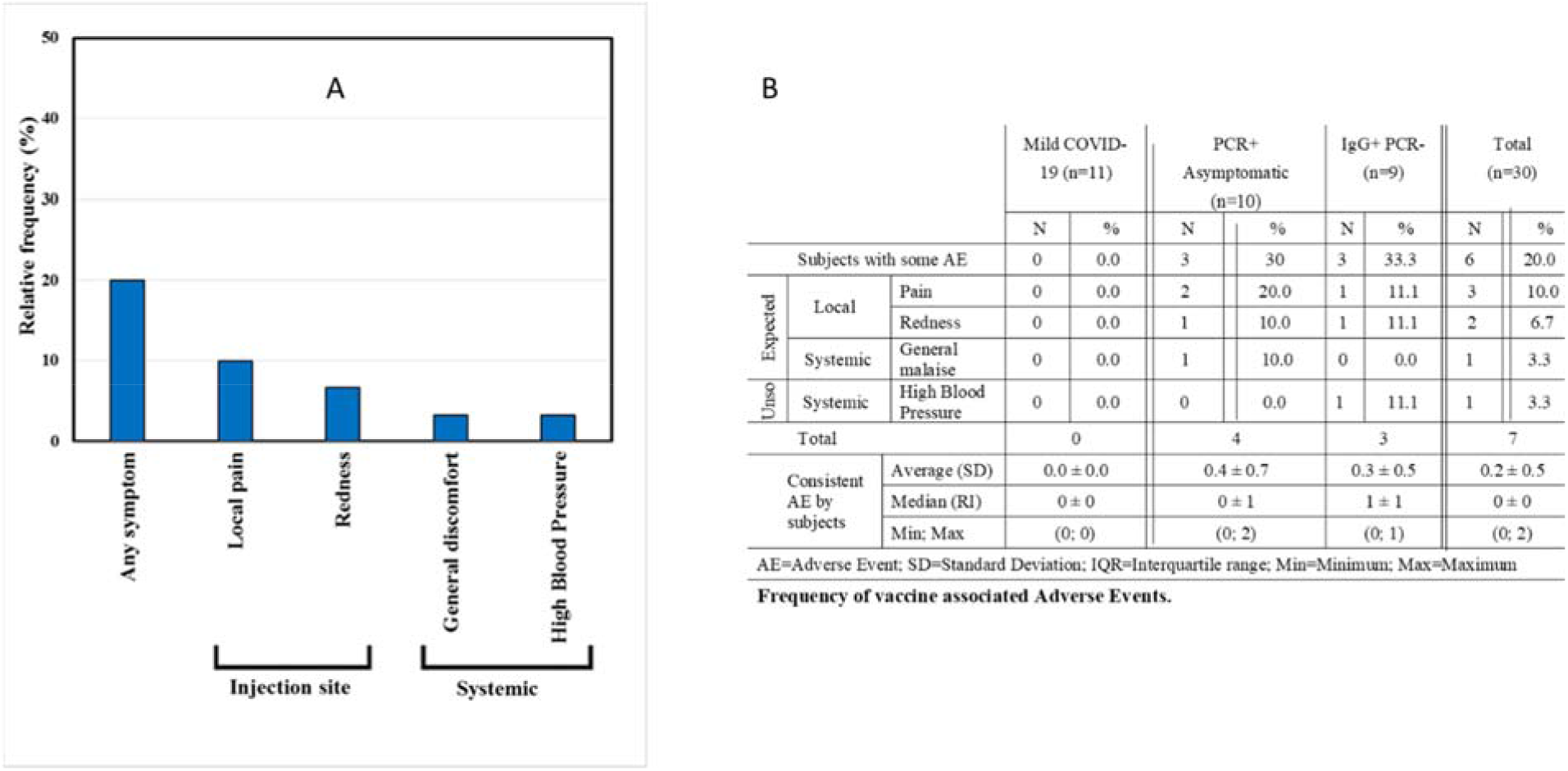
Vaccine-associated adverse events following immunization vaccination with a single dose of FINLAY-FR-1A (N= 30 individuals)

### Anti-RBD IgG antibody response

In 26 of 30 participants, a **s**ignificant increase of anti-RBD IgG was detected at post-vaccination day 7, showing evident stimulation of secondary antibody response, as previously reported in health care workers (HCW).^4,5^ IgG level further increased at day 14 and reached the highest point at day 28, with medians of 330.4 and 722.2 AU/mL respectively. Antibody level at day 28 represents a 21-fold increase compared to the pre-vaccination level and a 10-fold increase compared to a Cuban Convalescent Serum Panel (CCSP) (pre-vaccination: [N=30]; Median=34 AU/mL, and CCSP [N=47]; Median=67 AU/mL) (Figure 2 and Table 1). This result is similar to a previous finding with one dose of mRNA vaccine BNT162b2.^5^ An increase in anti-RBD IgG is observed in all three subgroups, with no differences between them (not shown). There were only four non-responders (NR, 13%) distributed in the three subgroups.

**Table 1:**
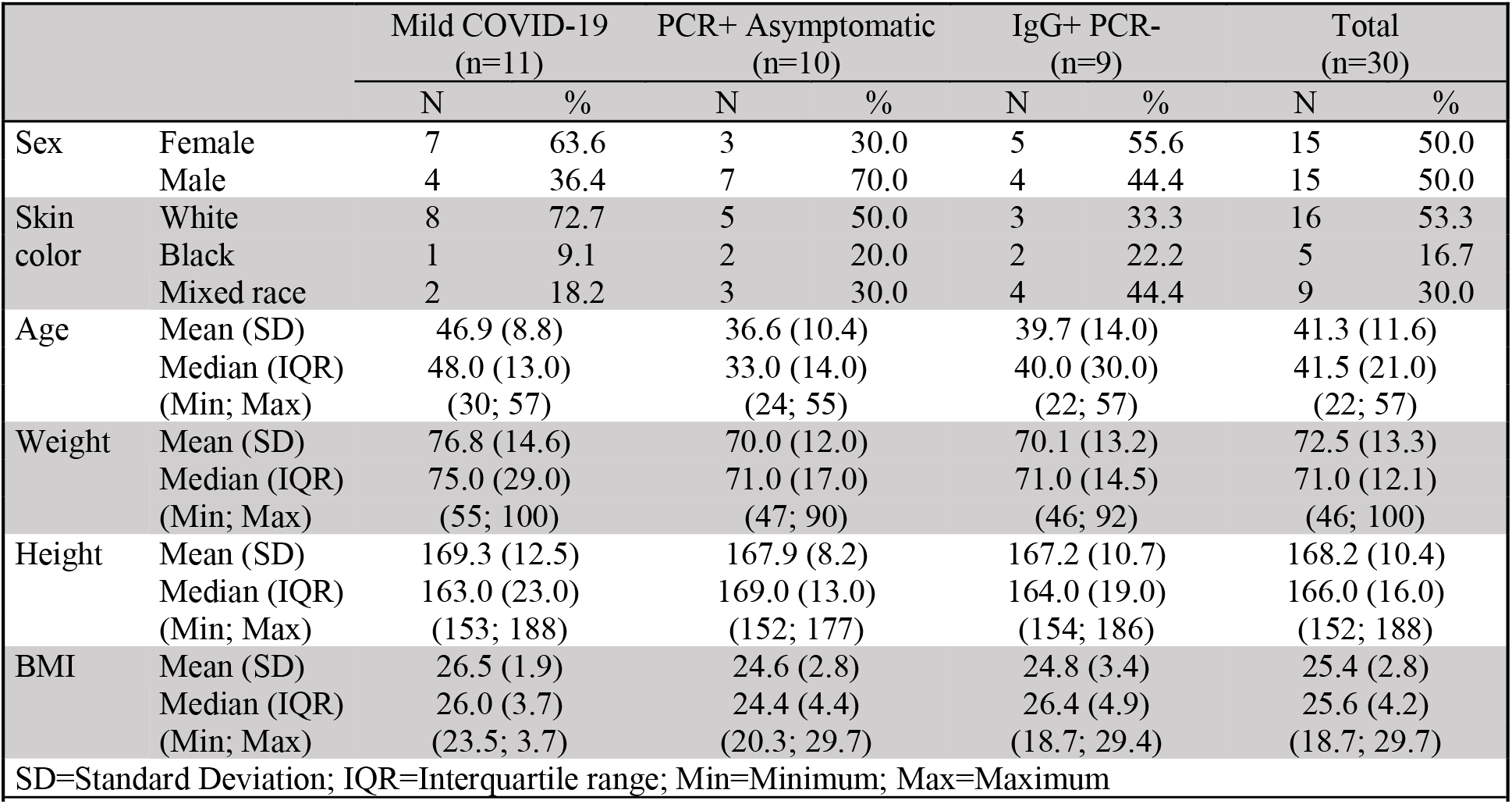
Demographic characteristics.

|  |  | Mild COVID-19<br>(n=11) |  | PCR+ Asymptomatic<br>(n=10) |  | IgG+ PCR-<br>(n=9) |  | Total<br>(n=30) |  |
| --- | --- | --- | --- | --- | --- | --- | --- | --- | --- |
|  |  | N | % | N | % | N | % | N | % |
| Sex | Female | 7 | 63.6 | 3 | 30.0 | 5 | 55.6 | 15 | 50.0 |
|  | Male | 4 | 36.4 | 7 | 70.0 | 4 | 44.4 | 15 | 50.0 |
| Skin color | White | 8 | 72.7 | 5 | 50.0 | 3 | 33.3 | 16 | 53.3 |
|  | Black | 1 | 9.1 | 2 | 20.0 | 2 | 22.2 | 5 | 16.7 |
|  | Mixed race | 2 | 18.2 | 3 | 30.0 | 4 | 44.4 | 9 | 30.0 |
| Age | Mean (SD) | 46.9 (8.8) |  | 36.6 (10.4) |  | 39.7 (14.0) |  | 41.3 (11.6) |  |
|  | Median (IQR) | 48.0 (13.0) |  | 33.0 (14.0) |  | 40.0 (30.0) |  | 41.5 (21.0) |  |
|  | (Min; Max) | (30; 57) |  | (24; 55) |  | (22; 57) |  | (22; 57) |  |
| Weight | Mean (SD) | 76.8 (14.6) |  | 70.0 (12.0) |  | 70.1 (13.2) |  | 72.5 (13.3) |  |
|  | Median (IQR) | 75.0 (29.0) |  | 71.0 (17.0) |  | 71.0 (14.5) |  | 71.0 (12.1) |  |
|  | (Min; Max) | (55; 100) |  | (47; 90) |  | (46; 92) |  | (46; 100) |  |
| Height | Mean (SD) | 169.3 (12.5) |  | 167.9 (8.2) |  | 167.2 (10.7) |  | 168.2 (10.4) |  |
|  | Median (IQR) | 163.0 (23.0) |  | 169.0 (13.0) |  | 164.0 (19.0) |  | 166.0 (16.0) |  |
|  | (Min; Max) | (153; 188) |  | (152; 177) |  | (154; 186) |  | (152; 188) |  |
| BMI | Mean (SD) | 26.5 (1.9) |  | 24.6 (2.8) |  | 24.8 (3.4) |  | 25.4 (2.8) |  |
|  | Median (IQR) | 26.0 (3.7) |  | 24.4 (4.4) |  | 26.4 (4.9) |  | 25.6 (4.2) |  |
|  | (Min; Max) | (23.5; 3.7) |  | (20.3; 29.7) |  | (18.7; 29.4) |  | (18.7; 29.7) |  |
| SD=Standard Deviation; IQR=Interquartile range; Min=Minimum; Max=Maximum |  |  |  |  |  |  |  |  |  |
| Table 1: Demographic characteristics. |  |  |  |  |  |  |  |  |  |

**Figure 2.**
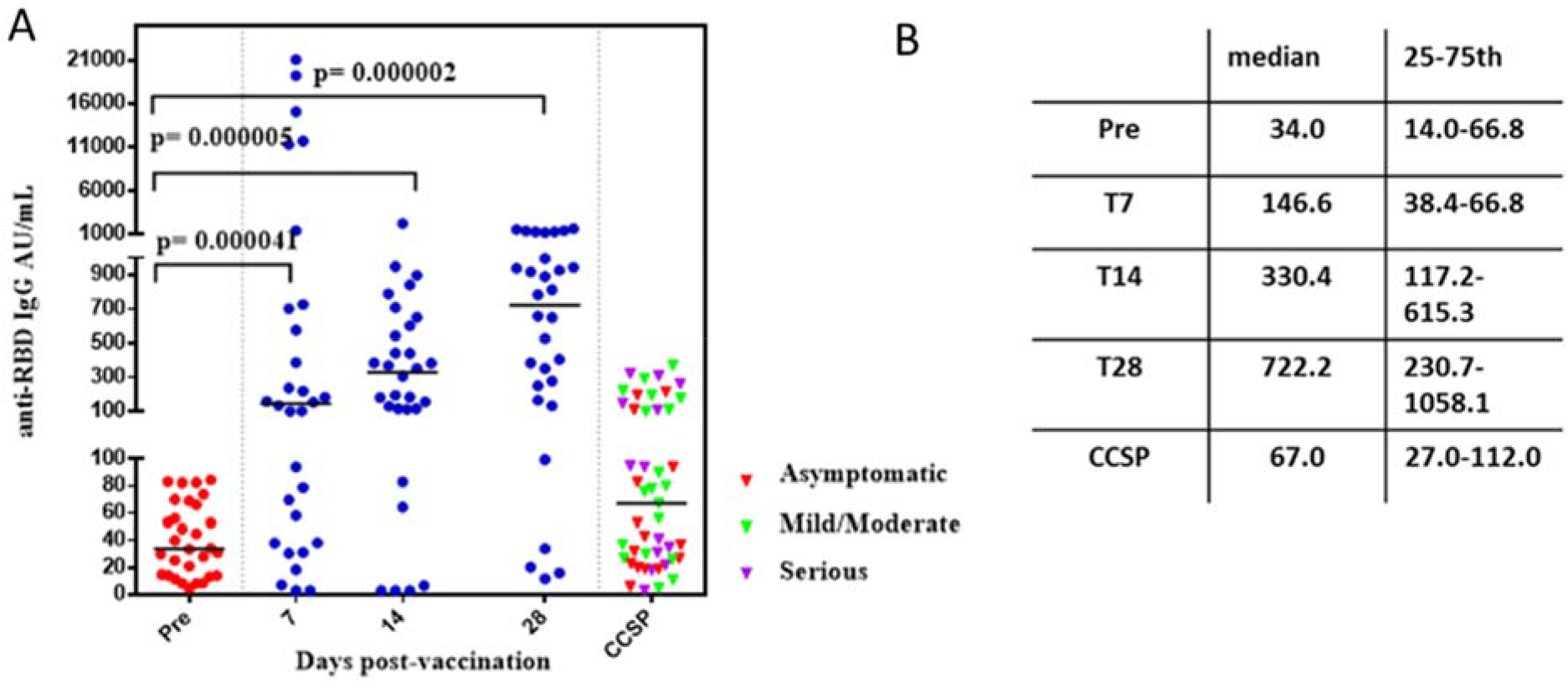
**A**. Immunogenicity after single dose of FINLAY-FR-1A vaccine in convalescents of SARS-CoV-2 (N=30). Anti-RBD IgG concentration at pre-vaccination and at 7 (T7), 14 (T14) and 28 (T28) days post-vaccination, expressed in arbitrary units/mL. CCSP: Cuban Convalescent Serum Panel (N=47). **B**. Median IgG and 25-75 percentile.

### Functionality of antibodies

The efficacy of anti-RBD antibodies in blocking interaction between recombinant RBD and hACE2 is evaluated in an ELISA inhibitory test.^6^ We studied quality of antibodies elicited by natural infection (before vaccination) and after single-dose vaccination. Interaction between recombinant RBD and recombinant hACE2 can be blocked by anti-RBD antibodies as a primary indicator of functionality, as determined by this type of ELISA test.^7^ We measured the inhibition ratio of RBD–hACE2 interaction at a serum dilution of 1/100 (Figure 3A) and the molecular virus neutralization titer (mVNT_50_) (Figure 3 B). mVNT_50_ is the serum dilution inhibiting 50% of RBD: hACE2 interaction.

**Figure 3.**
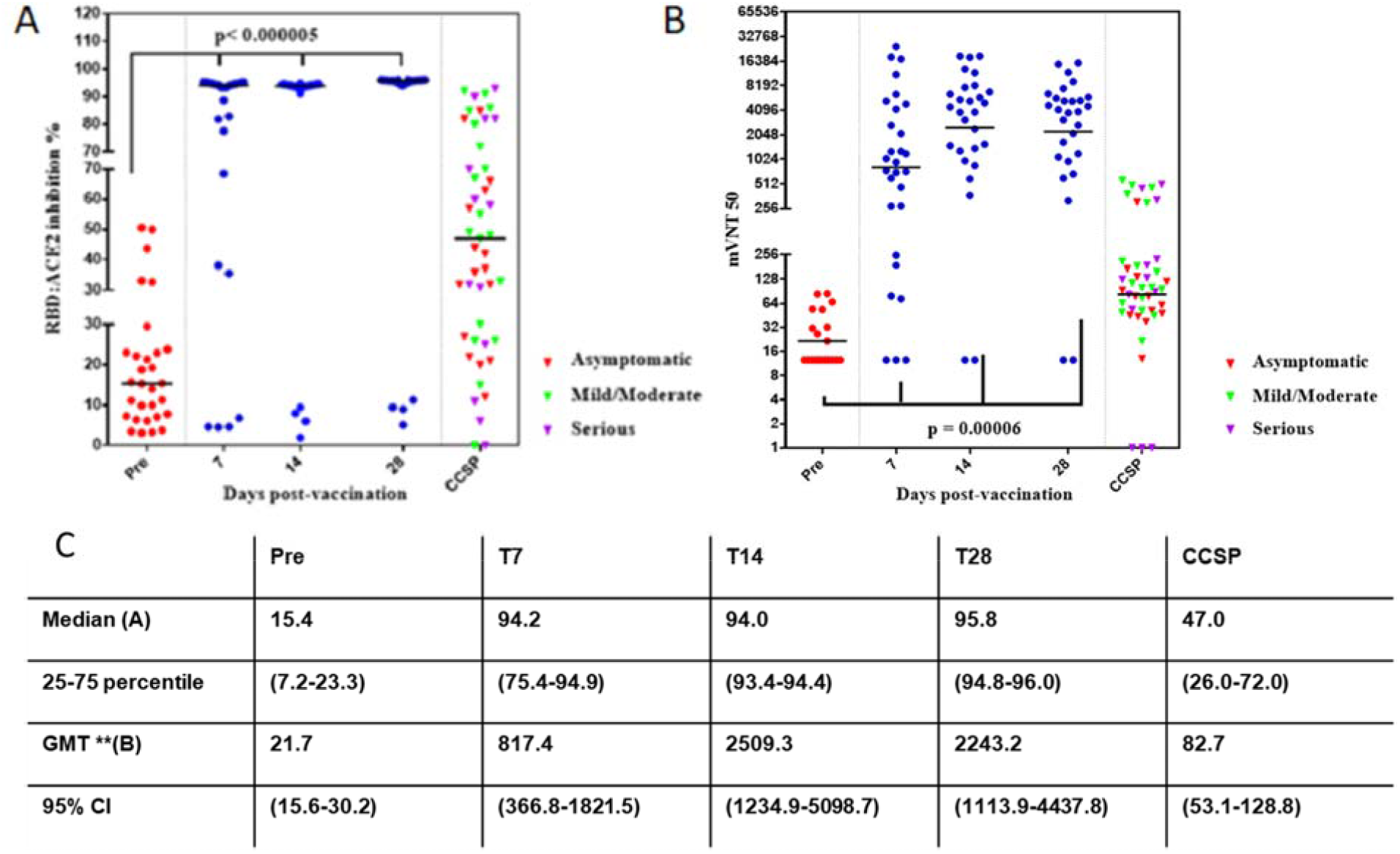
Anti-RBD IgG antibodies’ capacity for inhibiting RBD–hACE2 interaction, as measured by competitive ELISA. **A**) % Inhibition of RBD–hACE2 interaction at 1/100 serum dilution, median and 25-75percentile **B**) mVNT_50_ determining as the maximum serum dilution, given 50 % inhibition of RBD–hACE2 interaction. **C**) GMT** and 95 % CI do not consider NR.

**Figure 4.**
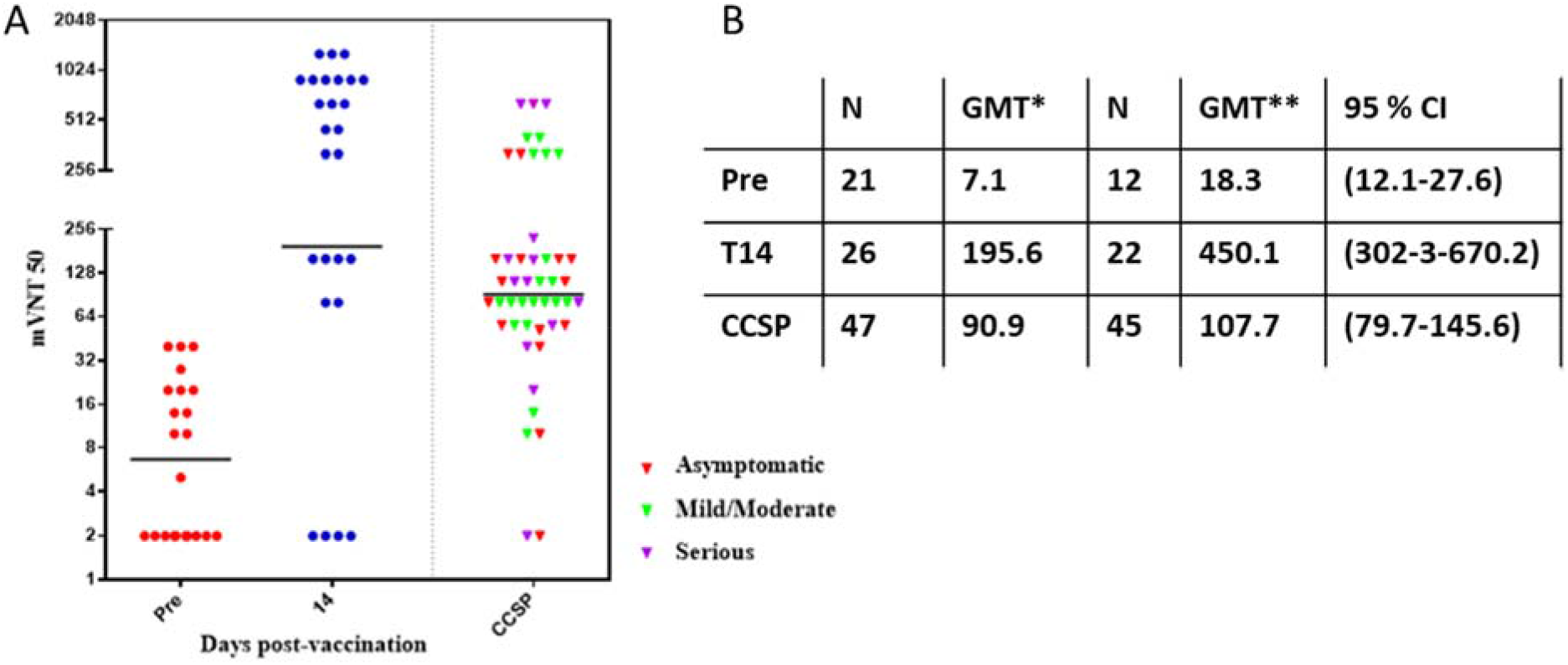
**A)** Virus neutralization Titer pre-vaccination, T14 after vaccination and CCSP including NR. **B**) The GMT* reported considers NR. GMT** and 95% CI do not consider NR.

For all study participants, the RBD: hACE2 inhibition ratio was below 60% before vaccination; after a single vaccine dose, this ratio increased over time for 26 of 30 participants (Figure 3A). All these responders (N=26; 87%) attained a 94% inhibition ratio:19 did so at day 7 post-vaccination and the remaining 7 did so at day 14. The CCSP showed a median of 47% inhibition.

To evaluate the functionality of antibodies, the mVNT_50_ value is an important complement to the information derived from the RBD: hACE2 inhibition ratio. The GMT** of mVNT_50_ at day 28 post-vaccination represents a 118-fold increase over the pre-vaccination value (2243.2/21.7) and a 27-fold (2243.2/82.7) increase over the CCSP value.

Finally, the conventional virus neutralization titer, (cVNT_50_) with live SARS-CoV-2 was evaluated 14 days after vaccination. A GMT** (not considering NR) obtained for vaccinated convalescents is cVNT_50_ = 450.1 (N=22), 4-fold compared to the median obtained for CCSP (cVNT_50_=107.7, N=47).

The results obtained from this phase I clinical trial demonstrated the safety and immunogenicity of a single dose application of FINLAY-FR-1A to SARS CoV-2 convalescent and pave the way for a phase II clinical trial to establish a general convalescent vaccination protocol.

## Data Availability

Data will b e available at https://rpcec.sld.cu/trials/RPCEC00000349 or under request to author

## Acknowledgment

We thank the individuals enrolled in the study for their generosity, as well as support from Dr. Lila Castellanos and Gail Reed for editorial assistance in English.

## Ethics statement

The clinical trial study protocol was reviewed and approved by the Ethics Committee of the Hematology and Immunology Institute in Havana, Cuba. All participants provided written informed consent prior to the participation. A detailed description is provided in the supplementary materials.

## Supplementary Information

### Ethical considerations

The Cuban Ministry of Public Health has established a medical care program for COVID-19 convalescent patients, aimed at prevention of further health problems, which provides rehabilitation services to this vulnerable group. The Hematology and Immunology Institute—clinical site of the trial—and the trial’s clinical research team are included in this medical care program.

The Cuban Ministry of Public Health and the Cuban National Regulatory Agency (Center for State Control of Medicines and Medical Devices, CECMED), approved this clinical trial carried out in COVID-19 convalescent individuals.

The clinical trial was conducted in accordance with the principles of the Declaration of Helsinki and Good Clinical Practice. Approval was obtained from the Independent Ethics Committee for Studies on Human Subjects, at the Hematology and Immunology Institute, Havana, Cuba. This Committee is structured and staffed appropriately; its seven members cover diverse expertise and it includes one community member. The Committee approved the vaccine used in the clinical trial as well as the procedures described in the protocol.

An Independent Data Monitoring Committee ensured respect for the interests and welfare of trial participants. This Committee comprises four members specializing in clinical trials and data monitoring, and is completely independent of the sponsor and clinical investigators. It performed an interim analysis of data at day 14 post-vaccination, with emphasis on safety and reactogenicity. Final monitoring of safety, reactogenicity and immunogenicity was performed 28 days post-vaccination.

During the study, both Independent Committees assessed the trial’s risk–benefit ratio and assured the rights, health and privacy of volunteers, including the confidentiality of information.

During recruitment, investigators provided potential participants with extensive relevant information, both oral and written. All questions and doubts were clarified. The decision to participate in the study was completely voluntary. Written informed consent was obtained from all participants.

## Methods

A phase I, open, adaptive and monocentric clinical trial was designed to evaluate the FINLAY-FR-01A vaccine against SARS-CoV-2. The study is ongoing and is registered at the Cuban Public Registry of Clinical Trials: RPCEC 00000349.^1^ Thirty convalescent individuals of mild COVID-19 and individuals with subclinical infections, aged 19–59 years, and with levels of inhibitory antibodies of RBD-ACE2 binding <60%, were included in three subgroups:

A: Recovered from mild COVID-19

B: PCR-positive, asymptomatic persons

C: Individuals with subclinical infection detected by community-based prevalence studies, seropositive with specific IgG but negative PCR.

Individuals included in subgroups A and B were admitted in hospital according to Cuban protocols for SARS-CoV2 infection. At study inclusion, they had been discharged fat least two months’ earlier.

Participants received a single intramuscular injection of the FINLAY-FR-01A vaccine at a dose of 50 µg dimeric-receptor binding domain (d-RBD) in aluminum hydroxide gel. Safety and reactogenicity evaluations were the primary outcome, assessed by occurrence of adverse events over 28 days post-vaccination. As secondary outcome, vaccine immunogenicity was evaluated. Humoral responses at baseline and following vaccination were evaluated using:

1. In-house standardized IgG ELISA against d-RBD
2. *In vitro* molecular neutralization test based on antibody-mediated blockage of hACE2– RBD interaction expressed either as % inhibition at a dilution of 1/100 or mVNT_50_ (the maximum dilution of the serum affording 50% inhibition).
3. Viral neutralization test.

In this preliminary report, we describe the safety, reactogenicity and immunogenicity of vaccination with 50 µg d-RBD in single-dose.

### Study design

This phase I, adaptive, open clinical trial was carried out at the Hematology and Immunology Institute in Havana, Cuba.

During the screening, a full medical history was recorded for each participant, and blood was drawn to test for: HIV, HCV, hepatitis B, VDRL, full blood count, kidney and liver function, pregnancy (through rapid test in women of childbearing age); to measure the background of IgG anti-RBD, blocking antibodies of ACE2-RBD interaction, viral neutralization test and to explore cellular immunity. PCR tests were done once between 7 days and 72 hours before inclusion.

Exclusion criteria were: severe COVID-19, any acute disease seven days before recruitment, non-controlled chronic diseases, primary or secondary immune deficiencies, history of severe allergy, pregnancy, breastfeeding, immunological treatment during prior 30 days, and hospitalization for COVID-19 during the two months before recruitment. An inhibition rate of RBD–hACE2 interaction >60% and SARS-CoV-2 PCR positive tests were also exclusion criteria. Forty COVID-19 convalescent individuals were recruited and thirty of them were included after applying selection criteria.

### Product under evaluation

FINLAY-FR-01A vaccine: d-RBD 50 µg absorbed in aluminum hydroxide gel (1.25 mg). The vaccine was manufactured according to current Good Manufacturing Practice by the Finlay Vaccine Institute in Havana, Cuba.

### Procedures

Convalescent individuals were recruited according to previously described criteria. The Independent Data Monitoring Committee and Independent Ethics Committee checked recruitment and vaccination proceedings. Volunteers were considered enrolled into the clinical trial at the moment of vaccination.

Convalescent individuals had blood samples extracted to evaluate safety at days 0 (pre-vaccination) and 28 (after vaccination). Blood samples for immunology studies were taken at days 0, 7, 14 and 28.

Volunteers were closely observed for 3 hours after vaccination. After vaccination, active surveillance was carried out at days 1, 2, 3, 7, 14 and 28. Passive surveillance was also done using diaries during the 28-day follow-up period.

Expected and protocol-defined local site reactions (injection site pain, warmth, redness, swelling, induration) and systemic symptoms (general malaise, rash, and fever defined as an axillary temperature of ≥38 °C) were recorded for 7 days. All other events were recorded for 28 days. Possible serious adverse events were carefully monitored throughout the follow-up period.

Severity of expected and protocol-defined local and systemic adverse events were graded as mild, moderate and severe, according to Brighton Collaboration definition and the Common Terminology Criteria for Adverse Events version 5.0.

Severity of unsolicited adverse events were graded with the following criteria: mild (transient or mild discomfort, no interference with activity), moderate (mild to moderate limitation in activity), severe (marked limitation in activity).

All adverse events were reviewed for causality by investigators, and events were classified according to WHO inconsistent causal association to immunization, consistent causal association to immunization, indeterminate, unclassifiable.^2^

Humoral immune response at baseline and following vaccination was evaluated by:

1. In-house quantitative IgG ELISA to detect antibodies against d-RBD, based on d-RBD as coating antigen. The assay uses an at-home standard characterized serum, which was arbitrarily assigned 200 U/mL (based on mVNT50 of 200 and cVNT of 160 for this serum). An anti-Human IgG-peroxidase conjugate was used for detection of anti-RBD IgG.
2. Molecular virus neutralization test is based on antibody-mediated blockage of RBD– hACE2 interaction. This test can be considered an *in-vitro* surrogate of virus neutralization test, detecting neutralizing antibodies in a short time. It uses recombinant RBD-mouse-Fc (RBD-Fcm) and the host-cell receptor hACE2-Fc (ACE2-Fch) as coating antigen. Human antibodies against RBD can block the RBD– Fcm interaction with its receptor. The RBD–Fcm that was not inhibited can bind to ACE2–Fch, and is recognized by a monoclonal antibody anti-gamma murine conjugated to alkaline phosphatase. This inhibition ELISA mimics the virus–host interaction at the molecular level.^3^
3. Viral neutralization test. This is the gold standard for determining antibody efficacy against SARS-CoV-2. It is a colorimetric assay based on antibody neutralization of SARS-CoV-2 cytopathic effect on Vero E6 cells was used^4^.

The viral neutralization test was used to evaluate samples before vaccination (0 day) and on day 14 after vaccination. The other humoral tests were applied at days 0, 7, 14 and 28 after vaccination.

A Cuban Convalescent Serum Panel (CCSP) well-characterized by standardized ELISA, *in vitro* inhibitory assay and viral neutralization test was used to evaluate vaccine-elicited immune response. It is composed of 47 serum samples from individuals recovered from mild, moderate and severe COVID-19.

### Statistical analysis

Calculation of the sample size was based on a serious adverse events rate lower than 5%. Two-sided 95% confidence intervals for one proportion were calculated, taking into account a target width of 0.194.

Safety and reactogenicity endpoints are described as frequencies (%). The mean, standard deviation, median, interquartile range, and the minimum and maximum values were used to describe the demographic characteristics and adverse events of the sample.

Median was calculated for immunological endpoints. For mVNT and cVNT, the geometric mean and 95% confidence intervals were calculated. Seroconversion rates for IgG antibodies anti-RBD (≥4-fold increase in antibody concentration over pre-immunization levels) were calculated, as well as frequency of participants with *in vitro* inhibitory antibody titers >70%.

Spearman’s rank correlation was used to assess relationships among techniques used to evaluate the humoral immune response. An alpha signification level of 0.05 was used. The Bayes Factor was used to carry out the risk-benefit analysis. The t Student’s t-test or the Wilcoxon Signed-Rank Test were used for before–after statistical comparison. Statistical analyses were done using SPSS version 25.0; EPIDAT version 4.1 and Prism GraphPad version 6.0.

## Results

Most convalescent individuals were of white skin color. Average age was 41 years, ranging from 22 to 57 years old, and body mass index of 25.4 kg/m^2^ (18.7 to 29.7 kg/m^2^). Half of all participants were women (Table 1).

The sample size calculation was based on a serious adverse event rate of less than 5%. No serious adverse events were reported in the clinical trial, so the sample size estimate was adequate (the probability of relating serious adverse events was 0.0322).

Mainly detected were mild adverse events. Local pain was the most common (10%) and redness (6.7%). Both were the only expected adverse events with consistent causal association to vaccination. The expected systemic reactions were limited to general malaise and mild fever, one subject each, with inconsistent causal association to vaccination (Figure 1). The severity and intensity of the local and systemic reactions was greatest on day one after vaccination and generally disappeared within the first three days.

Unsolicited adverse events were predominantly mild and moderate in nature and resolved during the follow-up period. The main unsolicited adverse event was high blood pressure, but only one case with consistent causal association to vaccination (3.3%). This subject was the only severe case in this clinical trial, but recovered within the first hours after vaccination. Volunteers with history of high blood pressure were admitted to the study if the pressure was controlled at recruitment.

